# SARS-CoV-2 antibody prevalence and symptoms in a local Austrian population

**DOI:** 10.1101/2020.11.03.20219121

**Authors:** Dennis Ladage, Yana Höglinger, Dorothee Ladage, Christoph Adler, Israfil Yalcin, Ralf J. Braun

## Abstract

**Background:** Since December 2019 the novel coronavirus (SARS-CoV-2) is the center of global attention due to its rapid transmission and toll on health care systems and global economy. Population-based serosurveys measuring antibodies for SARS-CoV-2 provide one method for estimating infection rates and monitoring the progression of the epidemic.

**Methods:** In June 2020 we succeeded in testing almost half of the population of an Austrian township (n=835 of 1359 inhabitants) with a reported higher incidence for COVID-19 infections. We determined the level of prevalence for SARS-CoV-2 in this population, factors affecting, and symptoms correlated with prior infection.

**Results:** We found a high prevalence of 9% positive antibodies among the town population in comparison to 6% of the neighboring villages. Only 20% of SARS-CoV-2 cases self-declared being asymptomatic. In contrast, we identified six single major symptoms, including anosmia/ageusia, weight loss, anorexia, general debility, dyspnea, and fever, and especially their combination to be of high prognostic value for predicting SARS-CoV-2 infection in a patient. Our comparison of the gold standard lab-based ELISA test and the on-site antibody test demonstrated a lack of accuracy for the latter test form.

**Conclusions:** This population study demonstrated a high prevalence of antibodies to SARS-CoV-2 as a marker of both active and past infections in an Austrian township. Several symptoms revealed a diagnostic value especially in combination. Results from self-administered antibody tests should be considered with caution.

---

The world is still in the midst of the coronavirus (SARS-CoV-2) pandemic with Austrian towns as Ischgl resembling as local epicenters. In June 2020 we succeeded in testing almost half of the population (47%) of an Austrian township with a reported higher incidence for COVID-19 infections. We determined the level of prevalence for SARS-CoV-2 in this population, factors affecting, and symptoms correlated with prior infection. Execution and design of study was in accordance with the local ethics committee and in approval by local and national authorities.

The township Weißenkirchen/Wachau (1359 inhabitants) is comprised of the town Weißenkirchen (926), and the communities Wösendorf (296), Joching (150), and St. Michael (23). People were recruited with a public call supported by local authorities as well as the Austrian red cross. 835 participants were tested for specific antibodies raised against SARS-CoV-2, with uniformly distributed sexes (48% male) and comprising people of all ages (ranging from 7 to 89). The participants completed a questionnaire on personal data as well as disease symptoms and their onset and duration.

Blood samples were split and used for detecting SARS-CoV-2-specific IgM and IgG antibodies in an onsite Rapid Test (WuHan UNscience Biotechnology) as well as in an certified diagnostic laboratory (Bioscientia, Ingelheim, Germany) utilizing enzyme-linked immunosorbent assay (ELISA). The reference method for screening and diagnosing Covid-19 infection is RT-PCR, nevertheless detection of antibodies against SARS-CoV-2 (IgG, IgM and IgA) play a complementary role with particular importance in providing epidemiological information. (1) Seroprevalence has been explored in Covid-19 patients confirmed by RT-PCR, as recently reviewed. (2) So far but only few studies have assessed seroprevalence in primarily asymptomatic individuals. Numbers were overall low even among high risk groups healthcare workers with frequent contact with Covid-19 patients ranging only at 1.6%. (3) In smaller studies in the general population rates only up to 5% were discovered. (4)

Using the onsite COVID-19 IgM/IgG Rapid Test, 3.4% (28/835) and 0.2% (2/835) of all tested subjects were shown to be positive for SARS-CoV-2 IgG and IgM, respectively. In contrast, in 8.5% (71/835) and 9.0% (75/835) of the tested subjects, SARS-CoV-2-specific IgG and IgA antibodies could be detected using the more sensitive and reliable laboratory-based ELISA assay. Only 31% (22/71) of the IgG positives detected with the ELISA test could also be determined by the IgG rapid test. Using the IgG ELISA data as the reference, we determined the test quality of the IgG rapid test as follows: sensitivity: 31%, specificity: 99%, positive predictive value: 79%, negative predictive value: 96%. The ELISA test is currently regarded as gold standard, therefore for the further analysis only those subjects that tested IgG ELISA positive were regarded as COVID-19 positive.

The high number of subjects with SARS-CoV-2-specific IgA antibodies could be a hint of more recent infections. (5) As the determination of IgA antibodies is far less reliable than for IgG, these data must be treated with caution, and were not considered in the following evaluation.

People, which developed SARS-CoV-2-specific IgG antibodies, stated significantly more often that they stayed abroad or in the Austrian state of Tyrol (42%, 30/71), as compared to the total tested population (26%, 206/806). Notably, the national hot spot Tyrol was not the source of the virus, but rather other countries, most of all Israel. 53% (10/19) of people, which visited Israel in early 2020 developed SARS-CoV-2-specific antibodies. Thus, the virus most likely was introduced from outside hotspots into the local population, where it proliferated.

9% (61/695) of the tested people from the township Weißenkirchen developed SARS-CoV-2- specific IgG antibodies, in contrast to 6% (10/167) in the control group of tested people from neighboring municipalities. Within the township Weißenkirchen, 10% (45/458) of the subjects from the town Weißenkirchen and 38% (6/16) from St. Michael, but only 5% (6/114) from Wösendorf and 6% (4/71) of Joching produced virus-specific antibodies. Thus, as expected, the township Weißenkirchen was more affected by SARS-CoV-2 as compared to neighboring municipalities, and that within the township the infection rates could be mainly localized to the town Weißenkirchen and the community St. Michael.

54% (38/71) of subjects with SARS-CoV-2 specific antibodies were male, as compared to 48% (404/834) in the total tested population. From our data a higher vulnerability of the male population as indicated by some studies is not evident. This however is in accordance with recent epidemiological data from Asia and especially large population analysis in China. (6) Similarly, we could not find a significant influence of age, body-mass index, or alcohol intake on the level of infection within the tested population.

Smokers turned out to be underrepresented upon subjects with SARS-CoV-2-specific antibodies. 8% of subjects with SARS-CoV-2 antibodies identified themselves as smokers, as compared to 17% in the total population. Since this observation did not reach statistic significancy, it remains open whether smoking may even reduce the risk of SARS-CoV-2 infections. Current data here suggest that smokers are more vulnerable and that smoking is an predictor for a negative outcome but not necessarily for a higher susceptibility to (asymptomatic) infection. (7)

20% (14/71) of all subjects which developed SARS-CoV-2-specific antibodies self-declared having noticed none of the 19 different disease symptoms listed in the questionnaire (Table 1). In contrast, 80% (57/71) of subjects with virus-specific antibodies self-declared one or more disease symptoms. Although some of these symptoms might be related to other diseases during the evaluation period, our data suggest that asymptomatic SARS-CoV-2 infections are rather uncharacteristic for the tested population. In fact, subjects with SARS-CoV-2-specific antibodies self-declared to have significantly more disease symptoms during the evaluation period than subjects lacking virus-specific antibodies (Table 1). Subjects having contact with SARS-CoV-2 demonstrated in a significant manner anosmia/ageusia, weight loss, anorexia, general debility, dyspnea, and fever, as compared to the total tested population. More and larger samples here will be necessary to confirm the prognostic values of symptoms found in other local studies. (8)

**Table 1:** Disease symptoms in the tested population. Data are from self-declaration of tested cases. Symptoms are ordered according to their enrichment in cases with SARS-CoV-2-specific antibodies (Asterisk * indicate significant enrichments)

| *Disease symptoms in the evaluation period (January to June 2020)* | *Number of total cases with symptoms* | *Number of SARS-CoV-2-positive cases with symptoms* |
| --- | --- | --- |
| Cases with any symptoms | 532 (63.7%) | 57 (80.3%) |
| Cases without any symptoms | 296 (35.4%) | 14 (19.7%) |
| Cases without data | 7 (0.8%) | 0 (0.0%) |
| *Anosmia/ageusia | 63 (7.5%) | 26 (36.6%) |
| *Weight loss | 33 (4.0%) | 11 (15.5%) |
| Apathy | 9 (1.1%) | 3 (4.2%) |
| *Anorexia | 49 (5.9%) | 15 (21.1%) |
| Pneumonia | 4 (0.5%) | 1 (1.4%) |
| *General debility | 147 (17.6%) | 24 (33.8%) |
| *Dyspnea | 51 (6.1%) | 8 (11.3%) |
| *Fever | 133 (15.9%) | 20 (28.2%) |
| Diarrhea | 105 (12.6%) | 14 (19.7%) |
| Stomachache | 60 (7.2%) | 7 (9.9%) |
| Headache / Pain in the limbs | 224 (26.8%) | 26 (36.6%) |
| Eczema | 21 (2.5%) | 2 (2.8%) |
| Tussis | 278 (33.3%) | 25 (35.2%) |
| Rhinitis | 301 (36.0%) | 27 (38.0%) |
| Somnolence | 12 (1.4%) | 1 (1.4%) |
| Sore throat | 222 (26.6%) | 16 (22.5%) |
| Swelling of the lymph node | 45 (5.4%) | 3 (4.2%) |
| Nausea / vomiting | 49 (5.9%) | 3 (4.2%) |
| Conjunctivitis | 28 (3.4%) | 1 (1.4%) |

In this community-based SARS-CoV-2 population study in Austria we found a high prevalence of 9% positive antibodies among the town population in comparison to 6% of the neighboring villages. Considering this large sample of almost half of the town population, we identified six single major symptoms and especially their combination to be of high prognostic value for predicting SARS-CoV-2 infection in a patient. Our comparison of the gold standard lab-based ELISA test and the on-site antibody test demonstrated a lack accuracy for this test form. Results from self-administered antibody test should be considered with caution. This study does have limitations, selection bias cannot be ruled out due to the volunteer nature of the study. Therefore, the estimated prevalence may be biased due to nonresponse or that symptomatic persons may have been more likely to participate. Ongoing additional population tests and follow up test regarding the prevalence in the town will provide further insights in the still developing and currently dynamic pandemic situation.

## Data Availability

All data for this manuscript is available.

## Notes

### Competing Interest Statement

The authors have declared no competing interest.

### Clinical Trial

Observational pilot study

### Funding Statement

No external funding to report.

### Author Declarations

Data was obtained observing national ethical guidelines. The study was reported to the ethics committee of Lower Austria (Niederoesterreich), by decision of the committee (letter from director of the board Robert Brucker) a ruling was waived due to the nature of the study.

## References

1. Yong SEF, Anderson DE, Wei WE, Pang J, Chia WN, Tan CW, et al. Connecting clusters of COVID-19: an epidemiological and serological investigation. Lancet Infect Dis. 2020;20(7):809–15.

2. Kontou PI, Braliou GG, Dimou NL, Nikolopoulos G, Bagos PG. Antibody Tests in Detecting SARS-CoV-2 Infection: A Meta-Analysis. Diagnostics (Basel). 2020;10(5).

3. Korth J, Wilde B, Dolff S, Anastasiou OE, Krawczyk A, Jahn M, et al. SARS-CoV-2-specific antibody detection in healthcare workers in Germany with direct contact to COVID-19 patients. J Clin Virol. 2020;128:104437.

4. Sood N, Simon P, Ebner P, Eichner D, Reynolds J, Bendavid E, et al. Seroprevalence of SARS-CoV-2-Specific Antibodies Among Adults in Los Angeles County, California, on April 10-11, 2020. JAMA. 2020;323(23):2425–7.

5. Ma H, Zeng W, He H, Zhao D, Jiang D, Zhou P, et al. Serum IgA, IgM, and IgG responses in COVID-19. Cell Mol Immunol. 2020;17(7):773–5.

6. Korean Society of Infectious D, Korean Society of Pediatric Infectious D, Korean Society of E, Korean Society for Antimicrobial T, Korean Society for Healthcare-associated Infection C, Prevention, et al. Report on the Epidemiological Features of Coronavirus Disease 2019 (COVID-19) Outbreak in the Republic of Korea from January 19 to March 2, 2020. J Korean Med Sci. 2020;35(10):e112.

7. Vardavas CI, Nikitara K. COVID-19 and smoking: A systematic review of the evidence. Tob Induc Dis. 2020;18:20.

8. Goyal P, Choi JJ, Pinheiro LC, Schenck EJ, Chen R, Jabri A, et al. Clinical Characteristics of Covid-19 in New York City. N Engl J Med. 2020;382(24):2372–4.

